# Limited Predictability of Client Attendance in a Support Program for HIV Vertical Transmission Prevention: A Comparison of Machine Learning and Community Health Worker Predictions

**DOI:** 10.1101/2025.08.26.25334431

**Authors:** Matthew Olckers, Alexander Lam, Lazola Makhupula, Mhlasakululeka Mvubu

## Abstract

Client attendance is vital for the success of HIV vertical transmission prevention programs, yet 23.4% of clients missed follow-up appointments after enrolling in a community health worker-led program (n=24,807, Aug-Dec 2022). Predicting which clients are most likely to miss appointments could enable targeted interventions to improve retention. While machine learning appears well-suited for this prediction task, its effectiveness compared to community health worker judgment remains unexplored. We evaluated three machine learning approaches—logistic regression, balanced random forest, and gradient-boosted trees—trained on client enrollment records (n=51,297 training; n=18,577 test) and compared their performance to predictions by community health workers (n=61), who possess direct client interactions and contextual insights. Machine learning models achieved modest predictive performance, with the balanced random forest showing the best accuracy (ROC AUC=0.689). Community health worker predictions similarly showed low accuracy despite their rich contextual knowledge, suggesting attendance is fundamentally difficult to predict. Qualitative insights identified complex and dynamic barriers including stigma, transportation difficulties, and competing personal commitments, often unpredictable at enrollment. These findings highlight fundamental limitations in predicting client attendance and suggest that merely accumulating additional data might not enhance predictive accuracy. Instead, resources might be better allocated to addressing systemic barriers identified by community health workers.

## 1 Introduction

Eastern and southern Africa carries a disproportionate burden of HIV, with approximately 55 percent of people and two-thirds of children living with HIV [31]. While parent-to-child (vertical) transmission of HIV can be effectively prevented through timely initiation and adherence to antiretroviral therapy, ensuring continuous engagement in care remains a major challenge. Missed appointments and interruptions in treatment undermine the success of prevention programs and threaten progress toward eliminating new pediatric infections.

To achieve positive health outcomes, providers of vertical transmission prevention (VTP) programs must ensure clients attend their appointments. If a provider knew which clients were most likely to miss appointments, they could target interventions and assistance to those clients. How can health providers predict which clients are most likely to miss their appointments?

As increasing amounts of health information are collected digitally, machine learning is a natural solution to predict client attendance, but most attempts have shown only modest accuracy [10]. The poor performance of machine learning to predict client attendance raises the question whether better data and models are needed, or attendance may be fundamentally difficult to predict.

We compare predictions generated by a machine learning model to predictions made by community health workers, who can gain rich, contextual, and nuanced information about their clients through direct interaction. The comparison of machine learning and community health worker predictions for the same clients provides deeper understanding of the limitations of machine learning than studying machine learning models in isolation.

We study these predictions in the setting of a VTP support program operated by mothers2mothers (m2m). Community health workers, which m2m call “mentor mothers”, support women living with HIV to start and adhere to treatment during pregnancy and breastfeeding. Clients meet with mentor mothers regularly, typically monthly or quarterly depending on the client’s circumstances. During the meetings, mentor mothers record information about their clients in a purpose-built mobile phone application so m2m can track client outcomes.

Studies on the m2m peer mentoring program have shown that it provides significant benefits to the mentees in terms of HIV prevention, diagnosis, and monitoring. Compared to mothers who were not enrolled in the m2m program, mentees were twice as likely to return for HIV testing (of both the mother and infant) after 6-8 weeks [29] and were significantly more likely to remain in care [3, 18], resulting in better well-being and VTP outcomes. However, these benefits require that clients adhere to the mentoring program, motivating our goal of predicting clients who are likely to miss appointments.

### Related literature

Since we compare two approaches for predicting client attendance—-machine learning and community health worker prediction—we contribute to two distinct research areas. First, we contribute to research that uses machine learning to predict whether clients attend health appointments.^1^ Second, we contribute to research that studies the accuracy of peer and community information.

Several studies have used machine learning to predict client attendance in healthcare settings, often with moderate success. A study on antenatal care attendance in Ethiopia, for instance, reported modest performance (ROC AUC 0.61–0.70) despite using rich health records [33]. In HIV programs, a rapidly growing body of work uses routine electronic records to predict missed visits or client retention. A common finding across these studies is the overwhelming predictive power of a client’s past attendance. In the United States, past missed visits were the strongest predictor of future attendance, with other clinical data adding little value [25], and even externally validated models using multi-site data achieved moderate AUCs of around 0.68, with retention history being a key feature [28]. Similarly, research in South Africa and Tanzania has shown that models using prior visit history and routine electronic data can effectively predict next-visit attendance and identify clients at high risk of disengagement [15, 16, 24]. To make our prediction task comparable between machine learning and the community health workers, we predict attendance using only information from a client’s first appointment, without the longitudinal histories central to prior models, yet we achieve comparable predictive performance.

On using local information to improve service delivery, a recent and growing literature shows that community members can hold valuable information about their peers. Examples include accurate knowledge of peers’ willingness to pay taxes [4], interest in buying land [23], business profitability [17], farming ability [22], and ability to spread information [5]. But even if community members are shown to have accurate information in one context, we cannot assume accurate knowledge in similar contexts. For example, Indonesian community members reported accurate wealth rankings about their peers [2] but a similar study, also in Indonesia, showed limited knowledge of fellow community members’ current income or need for financial support [30]. Researchers have found similar cases of inaccurate knowledge of neighbors’ poverty and wealth in Liberia [6] and Côte d’Ivoire [13]. Inaccurate knowledge extends to other domains besides wealth and poverty: in Peru, local leaders have been shown to have inaccurate knowledge of the incidence of intimate partner violence [1]. Our study documents inaccurate local knowledge in a new domain—the likelihood of attending health appointments—and complements the machine learning literature by providing a direct comparison between community predictions and algorithmic predictions.

## 2 Methods

This mixed-methods study examined client attendance in a VTP support program through two complementary approaches. First, we quantitatively evaluated and compared predictions of client attendance generated by machine learning models with predictions made by community health workers who had direct client interactions. Second, we conducted a thematic analysis of open-ended survey responses from community health workers to understand the underlying factors and barriers that influence client attendance. This mixed-methods approach allowed us to assess not only the predictive accuracy of different methods but also to explore why attendance may be inherently difficult to predict in this context.

This research project was granted ethical approval by UNSW Sydney’s Human Research Ethics Advisory Panel on 14 March 2023 (reference HC230085). The data utilized in this study originated from two sources managed by m2m: routine client records and a dedicated survey of community health workers. For the client data, which was collected during the registration process and throughout program participation; m2m obtained informed consent from these clients for the use of their data during enrollment. For the survey data, potential participants were provided with a detailed information sheet explaining the study’s purpose, data usage, and confidentiality measures. Each participating community health worker provided informed consent for the use of their survey responses prior to participation. After these internal data collection and consent procedures were completed by m2m, a de-identified version of the dataset was shared with the external research team for analysis, consistent with the ethical approval obtained (reference HC230085).

### 2.1 Machine learning approach

#### Data

We used de-identified data recorded at the client’s first m2m appointment. The m2m data includes clients based in Malawi, Uganda, Lesotho, South Africa, and Kenya.

The data was split chronologically for training (n=51,297 clients enrolled Jan-Oct 2021) and testing (n=18,577 clients enrolled Nov 2021-Jan 2022) to simulate real-world model deployment. This approach ensures that the model is trained on past data and then evaluated on its ability to predict outcomes for new clients who enroll in the future. By using a later time period for the test set, this method realistically assesses how the model would perform when deployed in a live setting, where it must make predictions for clients not seen during its training phase. The test set was used only to choose the best performing model, from a set of models that had already been tuned using a time series split on the training data.

A separate comparison set (n=24,807 clients enrolled Aug-Dec 2022) aligned with the timing of the community health worker survey. This specific cohort of clients is crucial as they were new to both the machine learning model and the community health workers at the time of the prediction task. The model, which was trained on data from Jan-Oct 2021 (and selected based on a performance on the test set of Nov 2021-Jan 2022 data), had no prior exposure to these clients. Concurrently, community health workers were surveyed to make predictions for their most recent new clients. This design allows for a direct comparison of predictive accuracy on the same set of clients.

### Target Measure

We target an indicator for whether a client missed their next scheduled appointment, defined as no attendance record within ±7 days of the scheduled date. For instance, if a client’s appointment was scheduled for November 15, the model would check for any attendance record for that client between November 8 and November 22. If the client visited the clinic on any day within this 15-day window, they would be marked as having attended. Conversely, if no visit was recorded during this entire period, they would be classified as having missed the appointment.

We focus on the first follow-up appointment after initial enrollment because it provides a consistent prediction task that is directly comparable between machine learning models and community health workers. At this stage, predictions can only draw on information collected during the enrollment visit, ensuring that neither approach relies on past attendance history, which is unavailable for new clients. By concentrating on the first follow-up, we capture a moment when targeted interventions can still be offered early in the client’s care journey.

Non-attendance rates were 25.9% (training), 26.8% (test), and 23.4% (comparison set). Figure 1 illustrates non-attendance rates over time, by plotting the share of clients that missed their first follow-up appointment in each month for which we have data. The share lies between 20 and 30 percent in most months. We had access only to the data necessary to train and test the machine-learning models (Jan 2021 - Jan 2022) and the data necessary to compare the machine-learning and community health worker predictions (Aug-Dec 2022), so there is a break in series between February and July 2022.

**Fig. 1.**
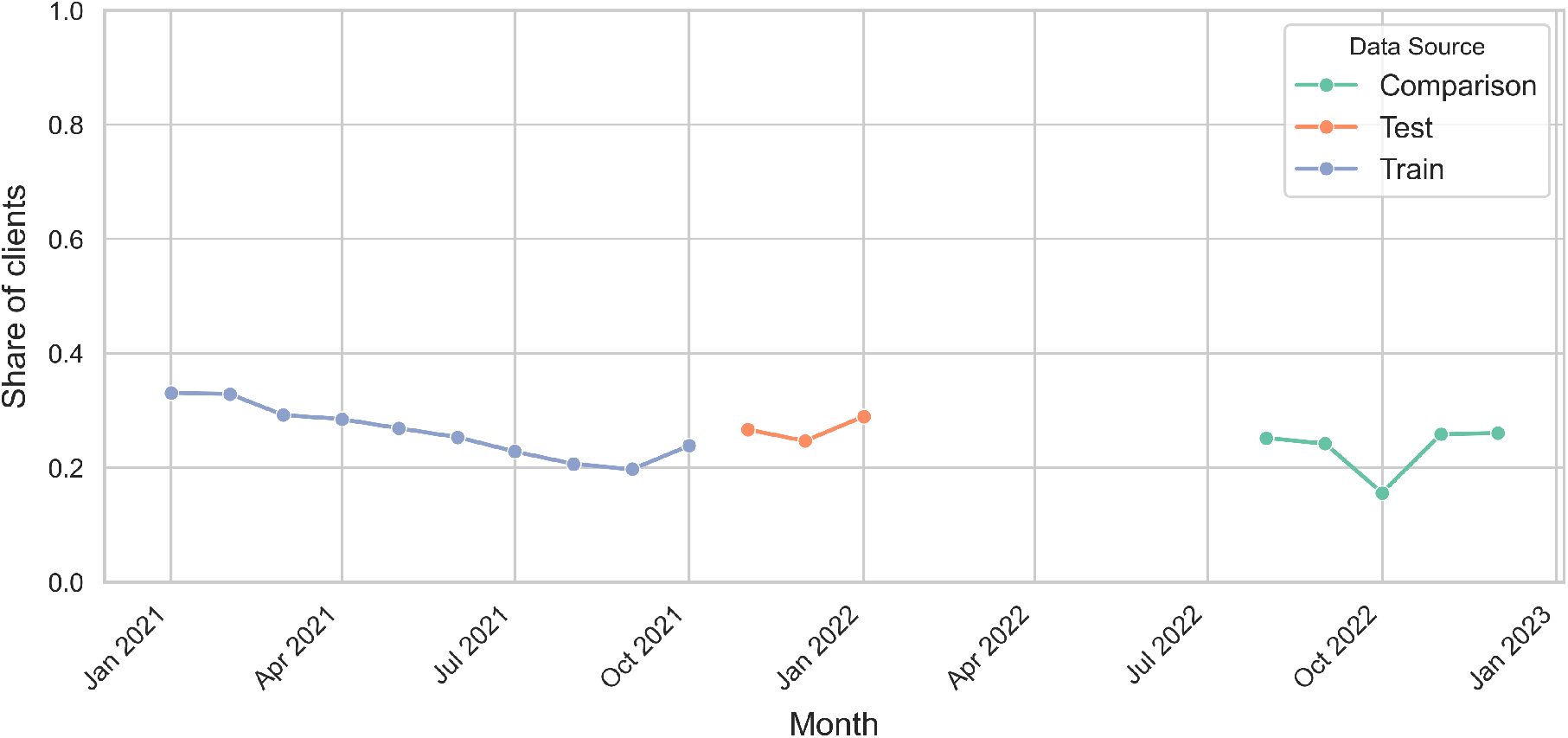
Share of clients that missed their first follow-up appointment

#### Features

Input features for machine learning models were restricted to data available at the initial enrollment meeting (Table 1), including demographics, program details, contact permissions, and basic HIV-related information. Data was captured by the community health workers using a mobile device.

**Table 1.** Selected Features.

| Feature | Mean |
| --- | --- |
| <u>Continuous features</u> |  |
| Age in years | 25.39 (8.14) |
| Duration to complete form in minutes | 6.83 (6.60) |
| Days to expected due date | 134.69 (56.80) |
| <u>Indicators (1 = Yes, 0 = No)</u> |  |
| m2m client before | 0.10 |
| Postnatal client | 0.18 |
| Know expected due date | 0.61 |
| Did not agree to follow up contact | 0.04 |
| Can contact with home visit | 0.75 |
| Can contact with SMS | 0.17 |
| Can contact with phone call | 0.74 |
| Own HIV status is positive | 0.30 |
| Own HIV status is unknown | 0.07 |
| Partner HIV status is positive | 0.16 |
| Partner HIV status is unknown | 0.33 |
| Knows own date of birth | 0.92 |
| Low or moderate adherence to treatment | 0.02 |

*Notes:* We report the mean of the feature. For continuous features, we report the standard deviation in parenthesis. The summary statistics are calculated using the 51,297 clients included in the training dataset. The days to the expected due date is calculated using only the 31,205 antenatal clients that know their due date.

#### Models

We evaluated regularized logistic regression, balanced random forests [11], and several algorithms that use gradient boosted trees, namely XGBoost [12], CatBoost [27], and LightGBM [20]. Hyperparameters were tuned using five-fold time-series cross-validation on the training set, optimizing for ROC AUC. The best-performing model on the held-out test set was selected.

### 2.2 Community health worker predictions

To compare the machine learning predictions, we surveyed m2m community health workers between August and December 2022. The survey included two types of questions designed to capture community health workers’ predictions about client attendance. First, survey respondents were asked about their most recent new client and how likely they thought that client was to miss the next appointment. Second, they were asked to nominate, from all the new clients they had seen during the week of the survey, the one they believed was most likely to miss their next appointment.

For the most recent new client questions, the survey asked: “Think of your most recent new client. A new client is a client who is accessing m2m services for the first time. Please enter their first name, last name, and date of birth.” The survey then asked “Do you think {recent client name} will miss their next appointment?” and responses were recorded using a five-point Likert scale (‘definitely not’, ‘probably not’, ‘might or might not’, ‘probably yes’, ‘definitely yes’).

To assess whether higher community health worker Likert ratings (“definitely not” to “definitely yes”) were monotonically associated with a higher probability of missing the next appointment, we used the Cochran–Armitage linear-by-linear trend test. Because the matched Likert sample was small (n=61) and the upper categories were sparse (including zero misses in the highest categories), we computed a Monte Carlo permutation p-value conditional on the margins (fixed group sizes and total number of misses), which is valid under sparsity and avoids reliance on large-sample normal approximations. The alternative was prespecified as one-sided (increasing), reflecting the hypothesis that higher community health worker concern implies greater miss risk. The one sided test increases statistical power.

The nomination questions asked “How many new clients did you see this week?” and then followed with “Of the new clients you saw this week, which client do you think is most likely to miss their next appointment? Please enter their first name, last name, and date of birth.”

To compare missing rates between clients nominated by community health workers and those not nominated, while accounting for differences across workers, we analyzed 2 × 2 tables stratified by community health worker (nominated vs. not nominated × missed vs. attended). First, we combined the stratum-specific tables to estimate a single (“common”) odds ratio using the Mantel–Haenszel method. Next, we used the Cochran–Mantel–Haenszel (CMH) chi-square test to assess whether this pooled odds ratio differed from 1 (no association). Because the CMH test conditions on each stratum’s fixed row and column totals, strata with no within-stratum variation in exposure or outcome contribute no information and are effectively dropped. We also assessed whether effects varied across strata using the Breslow–Day test.

### 2.3 Qualitative data collection and analysis

The survey collected open-ended responses to two questions. Firstly, after the community health worker rated if they thought their most recent new client would miss their next appointment, the survey asked: “What factors might prevent {recent client name} from attending their appointments?” Secondly, the final question of the survey asked: “You have reached the end of the survey. If you like, please share any thoughts or suggestions with the research team.” Although qualitative data is most commonly associated with interviews and focus groups with a small number of respondents, online surveys have the advantage of more easily reaching a large number of respondents, which allows for a ‘wide-angle lens’ on a topic [9]. We received 168 responses to the first open-ended question and 81 responses to the second.

We analyzed the survey responses using reflexive thematic analysis [7, 8], which is a qualitative research method that focuses on identifying themes and patterns within data while embracing the researcher’s insights. Rather than trying to separate the researcher’s biases and assumptions from the analysis, this approach values the researcher’s role in shaping the analysis.

We have used reflexive thematic analysis to benefit from the experience of our research team. Authors 3 and 4 worked for mothers2mothers and have extensive experience designing the mentor mother program. Reflexive thematic analysis allowed us to use insights from Authors 3 and 4 to improve our understanding of the survey responses. We use the following process to generate our themes:

- Author 1 read the survey responses several times and assigned initial codes. He used the codes to derive candidate themes.
- Author 1 then met with Authors 3 and 4 for feedback on the candidate themes, the codes, and selected quotes. Authors 3 and 4 shared insights to enrich the analysis.
- Author 1 shared a draft write-up of the analysis with Authors 3 and 4 for further comments. After incorporating the comments, Author 1 reread the survey responses and edited the draft.

## 3 Results

### 3.1 Machine learning predictions

We report the performance of the machine learning models in Table 2. The balanced random forest model performed best with an ROC AUC of 0.689, but still demonstrated limited ability to distinguish between clients who would attend versus miss their next appointment. Performance was comparable to similar studies [28, 33], even though these studies used many more predictors.^2^

**Table 2.** ROC AUC on the held-out test set by model.

| Model | ROC AUC |
| --- | --- |
| Logistic Regression | 0.598 |
| Balanced Random Forest | 0.689 |
| XGBoost | 0.687 |
| CatBoost | 0.682 |
| LightGBM | 0.681 |

*Notes:* ROC AUC values are calculated on the held-out test set. All models were tuned via randomized search with five-fold time-series cross-validation on the training set. The selected hyperparameters for each model are reported in Appendix E.

The ROC AUC captures the probability that the model assigns a higher risk score to a randomly chosen client who misses an appointment than to one who attends. A value of 0.5 corresponds to random guessing, while 1.0 indicates perfect classification. An AUC of 0.689 therefore means that in 68.9% of such pairs, the model correctly ranks the non-attending client higher than the attending client. Although this performance is clearly above chance, it also reveals substantial overlap in predicted risk scores between attenders and non-attenders, limiting the usefulness of the model for precise targeting.

Because this analysis is exploratory, m2m has not specified a particular classification threshold. Any threshold would need to be chosen with reference to the type and cost of the intervention being considered. In the absence of a defined intervention strategy, our aim here is simply to assess whether predictions based on enrollment data could, in principle, reach levels of accuracy that would justify targeted outreach.

### 3.2 Community health worker predictions

Each community health worker used a five-point Likert scale to record the likelihood of their client missing their next appointment. In total, 188 survey responses were collected, with the distribution across categories shown below:

- definitely not (71)
- probably not (50)
- might or might not (44)
- probably yes (16)
- definitely yes (7)

Even though 23.4 percent of clients missed their first appointment in the comparison set (which corresponds to the same period as the survey responses), only 12.2 percent of community health worker forecasts fell into the “probably yes” or “definitely yes” categories. However, if the middle category (“might or might not”) is interpreted as a 50% probability of missing, then the weighted average forecast across all 188 responses is 23.9%, remarkably close to the observed base rate. This suggests that while few workers were willing to classify clients as high risk, their aggregate predictions were reasonably well calibrated. However, calibration to the base rate provides only a partial picture; assessing predictive accuracy requires linking survey responses to client attendance records.

To test the accuracy of the community health workers’ predictions, we needed to match the name and date of birth of the client mentioned in the survey to m2m’s administrative data. Due to differences in spelling and the large number of clients with similar names, we were only able to match 61 clients to their administrative records. We took a conservative approach to ensure that, among the matched set, the attendance records corresponded to the correct client. The downside of this conservative approach is that our sample size was reduced from 188 to 61.

We adopted this conservative matching strategy to minimize the risk of linking a community health worker’s response to the wrong client record. Incorrect matches would artificially bias the predictive accuracy of community health worker judgments downwards, since outcomes would be evaluated against the wrong client. Although this approach reduced our matched sample size, it maximized the validity of the comparison by ensuring that only confidently linked responses were included. In practice, integrating such prediction questions directly into the mobile health application used by community health workers would remove the need for post-hoc matching and avoid these trade-offs, but developing and deploying new application features was beyond the scope of this project. Despite the smaller sample, the matched dataset provided enough observations to test whether community health worker judgments showed very high levels of predictive accuracy. However, we lack strong statistical power to detect more modest associations, so these results should be interpreted as indicative rather than definitive.

The share of clients who did miss their appointment, grouped by the five survey responses, is shown below. We also report the frequency of the response in brackets.

- definitely not (30): 10.0%
- probably not (19): 31.6%
- might or might not (9): 33.3%
- probably yes (2): 0%
- definitely yes (1): 0%

Of the community health workers in the matched set who selected that their client would definitely not miss their next appointment, 10% did miss their appointment. As soon as the community health worker indicated some uncertainty, by selecting ‘probably not’ or ‘might or might not’, the share of missing clients increases to just over 30 percent. Only three community health workers in the matched set selected ‘probably yes’ or ‘definitely yes’, and none of these three clients missed their next appointment.

Applied to the five-category Likert ratings, the Cochran–Armitage trend test showed a positive but non-significant association between higher community health worker concern and non-attendance: the permutation (Monte Carlo) p-value was 0.205 (one-sided, increasing), and the asymptotic linear-by-linear statistic yielded Z=0.954 with a corresponding one-sided p=0.170. We therefore do not reject the null of equal miss probabilities across Likert categories. The lack of statistical significance is consistent with the descriptive category counts—particularly the rarity of the highest categories and the absence of events in them, which limit power to detect a monotonic trend.

The survey also asked:

*Of the new clients you saw this week, which client do you think is most likely to miss their next appointment?*

We compare attendance rates for clients picked by the community health workers against the clients they did not pick. We matched 13 community health workers between the survey and administrative data who answered this question. We could only match a small number of community health workers because we needed to match both the username of the community health worker and the name of the client they nominated. The administrative data showed that these 13 community health workers saw a further 60 clients that they did not pick in the same week. For the 13 clients the community health workers picked, 15.4% missed their next appointment. For the 60 clients the community health workers did not pick, 15.0% missed their next appointment.

For 7 of the 13 community health workers, none of their clients missed their next appointment in the matched week. Therefore, these strata contained no within-community health worker outcome variation and were excluded by the conditioning step of the Cochran–Mantel–Haenszel (CMH) test. Among the remaining 6 informative strata, the CMH test provided no evidence that nomination status was associated with missing the next appointment (CMH *χ*^2^ = 0.061, *p* = 0.8045). The stratum-adjusted common odds ratio comparing nominated to non-nominated clients was 0.794 (95% CI 0.129–4.909), with a Breslow–Day test indicating no meaningful heterogeneity across workers (*χ*^2^ = 6.940, *p* = 0.2251). These results are consistent with the similar raw miss rates reported above (15.4% vs. 15.0%).

### 3.3 Comparison between machine learning and community health worker predictions

We can compare the accuracy of the machine learning predictions to the community health worker predictions for the 61 clients which the community health workers rated on the Likert scale, and for the set of 73 clients we linked to survey responses from 13 community health workers who were asked to pick which of their recent clients was most likely to miss their next appointment.

Each method could be used by m2m to create a targeting rule. For the machine learning approach, m2m could target the top k clients with the highest predicted likelihood of missing the next appointment. For the Likert survey question, m2m could target the clients who were above a threshold in the response. For example, they could target clients who the community health worker reports will ‘probably’ or ‘definitely’ miss their next appointment. We can compare machine learning and community health worker predictions by choosing each rule so that they target the same number of clients.

In Table 3, we simulate a targeting rule that targets a client if the community health worker reports that their likelihood of missing their next appointment is above a threshold. For example, in the third row of Table 3, the threshold is ‘might or might not’, so the rule would target clients for whom the community health worker reports as ‘might or might not’, probably yes’, or ‘definitely yes’ to miss their next appointment. In our sample of 61 clients, 12 clients would satisfy this rule. To compare the community health worker rule to a corresponding rule for machine learning, we take the top k clients with the highest predicted score from the machine learning model, where k is set to match the number of clients selected by the community health worker rule. So, for the third row, k = 12. Table 3 shows that the machine learning model has higher accuracy for predicting which clients will miss their next appointment.

**Table 3.** Comparison between machine learning (ML) and community health worker (CHW) predictions.

| Threshold | Top k | Accuracy % |  |
| --- | --- | --- | --- |
|  |  | CHW | ML |
| Definitely yes | 1 | 0 | 0 |
| Probably yes | 3 | 0 | 33.3 |
| Might or might not | 12 | 25.0 | 41.7 |
| Probably not | 31 | 29.0 | 35.5 |

*Notes:* Each row compares a targeting rule based on community health worker (CHW) predictions to an equivalent rule based on the machine learning (ML) model. For the CHW rule, the threshold indicates the minimum Likert category used to classify a client as high risk of missing their next appointment. For the ML rule, the number of targeted clients ( $k$ ) is set to match the number selected under the CHW threshold, and those with the highest predicted scores are chosen. “Top $k$ ” shows how many clients are targeted under each rule, and “Accuracy %” reports the share of these targeted clients who actually missed their next appointment.

We can perform a similar exercise to compare the question where the community health worker was asked to pick a client. Two of the 13 clients the community health workers picked (from a set of 73 clients) missed their next appointment. Among the top 13 clients (from this same set of 73 clients) with the largest scores predicted by the machine learning model, three missed their next appointment.

Although the machine learning approach was more accurate than the community health worker predictions, the difference is small.

### 3.4 Qualitative analysis of open-ended survey questions

Analysis of the open-ended survey responses revealed four main themes explaining client non-attendance: reluctance to attend, competing commitments, transport difficulties, and attending at alternative times or locations.

1. *Reluctance to Attend*: Some clients did not wish to attend appointments due to HIV-related stigma, concerns about medication availability at facilities, or the need for food before taking medication. Stigma was a particular concern for newly diagnosed clients; as one community health worker noted, her client might not attend because of “stigma because she’s newly tested in this month”. Lack of food could also lead to non-attendance, with one respondent explaining, “3 out of 10 clients in my caseload feels that they can’t take their drugs without food hence they opt to default on care”.
2. *Competing Commitments*: Clients often faced other important obligations that clashed with appointments. These included childcare responsibilities, work or study schedules, travel for work, and attending funerals. Work conflicts were highlighted: one client had “tight schedules e.g. work and not been given permission to attend her clinics”, while another “is a young mother who is still studying at high school. She might miss appointments because she might be busy at school. Nobody to take infant to facility”. Work-related travel could also interfere: one client is “a businesswoman and travels to the nearby country Mozambique and mostly misses appointments”.
3. *Transport Barriers*: Transport was the most frequently cited barrier. Challenges included distance, cost, lack of public transport, weather conditions, and reliance on partners for transport or funds. Lack of partner support could also be a significant issue: “The husband does not allow the wife to visit the health facility in general. For this time, she had to escape to come for the antenatal service”. Some clients also intentionally chose distant facilities due to stigma, compounding transport issues.
4. *Attending at Different Times or Locations*: Non-attendance at a scheduled m2m appointment did not always mean missing care altogether. Clients sometimes visit the clinic at a different time than scheduled to combine visits with partners or to coincide with other errands like collecting social grants. For instance, one client “has different days for child’s CPT and her refill dates hence might miss one to combine the two”, and another “waits for husband’s date of refill to come back to the clinic”. Clients might also attend other, non-m2m facilities, sometimes due to being walk-ins or traveling for work. As respondents noted: “Most of the clients who mostly misses appointment are walk-in from different facilities”, and another client “works in Kenya. And takes a lot of months to return, but she updates us when she gets refills from that side”.

These qualitative insights highlight factors, such as transport access or unexpected events like funerals, that are often dynamic, difficult to capture in administrative data available at enrollment, and thus hard to predict using either machine learning or community health worker judgment.

## 4 Discussion

This study highlights the difficulty in predicting client attendance at a VTP program based on initial enrollment data. Machine learning models showed modest performance (ROC AUC 0.689). Community health workers, who may possess rich contextual knowledge from client interactions, were also unable to predict attendance accurately.

The comparison between machine learning and community health worker predictions is a key contribution. The poor performance of community health workers suggests that the challenge lies not merely in the limitations of the features available to the machine learning algorithm, but in the fundamental unpredictability of the behavior itself in this setting.^3^ The information needed to predict attendance may be unknowable at the time of prediction.

Our qualitative findings corroborate this interpretation. Community health workers described a multitude of reasons for non-attendance related to stigma [26], competing commitments (work, childcare, funerals), transport barriers (cost, availability, weather, partner support), and complex healthcare navigation choices.^4^ Many of these factors are volatile, arise unexpectedly after enrollment, or depend on circumstances (like food insecurity or partner disapproval) that are difficult for community health workers, let alone algorithms, to ascertain reliably. This inherent complexity and dynamism likely places an upper bound on the achievable accuracy of any predictive model based on health care records.

While machine learning can identify some patterns associated with non-attendance, its utility for reliably targeting interventions in this VTP program appears limited. The modest accuracy achieved means that any targeting strategy based on these predictions would likely result in many false positives (intervening unnecessarily) and false negatives (missing clients who do need support).

These findings caution against an over-reliance on predictive analytics in complex public health settings. The assumption that better data or more sophisticated algorithms will inevitably lead to effective prediction may not hold when the target behavior is driven by highly dynamic, contextual, and often unobservable factors. Ethical considerations also arise; deploying potentially inaccurate or biased predictive models for targeting could lead to inefficient resource allocation, stigmatization, or failure to address the true, diverse needs of the client population [32].

Instead of focusing primarily on prediction, our results suggest that resources may be more effectively directed towards understanding and mitigating the systemic barriers to attendance identified through qualitative insights from community health workers and clients. Addressing issues like transport costs, inflexible clinic hours, stigma reduction, and ensuring consistent service quality could potentially improve attendance more broadly and equitably than relying on imperfect predictive targeting.

## Acknowledgments

We thank the mothers2mothers community health workers who participated in the survey and shared their valuable insights. We thank Mohammed Fadul, Garrit Gerke, Nakululombe Kwendeni, Raymond Moholisa, Moeti Moleko, and Robynn Paulsen-Naidoo from m2m for their assistance in preparing the data and their advice on m2m’s systems. We thank Tuba Islam from Google for her advice on machine learning approaches. We thank Brendan Maughan-Brown, Bryan Wilder and Lily Xu for helpful comments.

## B Authors’ contributions

MO designed the study and survey instrument in consultation with LM and MM. With support from their colleagues at m2m, LM and MM organized the health records used for machine learning and conducted the survey. MO and AL conducted quantitative data analysis and model training. MO completed the analysis of the qualitative data with input from LM and MM. MO wrote the manuscript with input from AL, LM, and MM.

## D Data Availability Statement

The data that supports the findings of this study are available from mothers2mothers. Restrictions apply to the availability of these data, which were used under license for this study. Data are available from the author(s) with the permission of mothers2mothers. The code used to replicate the results is available in the following repository: https://github.com/matthewolckers/m2m-predict-attendance

## E Hyperparameters

**Table 4.** Selected hyperparameters for best models (library parameter names)

| Model | Selected hyperparameters |
| --- | --- |
| Logistic Regression | `solver = lbfgs; penalty = l2; C = 0.0018329807108324356; max_iter = 1000` |
| Balanced Random Forest | `n_estimators = 400; max_depth = 10; min_samples_split = 2; min_samples_leaf = 4; bootstrap = True; replacement = True; sampling_strategy = "all"` |
| XGBoost | `n_estimators = 500; max_depth = 5; learning_rate = 0.01; subsample = 0.6; colsample_bytree = 0.9; gamma = 0.1; reg_alpha = 0.5; reg_lambda = 1; min_child_weight = 1` |
| CatBoost | `max_depth = 4; learning_rate = 0.05; l2_leaf_reg = 0.2; colsample_bylevel = 0.3; min_child_samples = 1; verbose = False` |
| LightGBM | `n_estimators = 200; max_depth = 4; num_leaves = 10; learning_rate = 0.05; colsample_bytree = 1; reg_lambda = 0.2; verbose = -1` |

*Notes:* Hyperparameters shown are for the selected best estimator determined via grid search using a time series split, with five splits. Unlisted parameters were left at library defaults. Replication code is available at: https://github.com/matthewolckers/m2m-predict-attendance

## Footnotes

1 We focus on studies that predict appointment attendance or retention in care. A related literature predicts medication adherence [19, 21].

2 One recent study reported an ROC AUC of 0.895 [14], but the study does not explain whether a held-out test set was used to measure performance. Measuring performance on the training data can inflate performance measures.

3 An alternative interpretation is that community health workers observed relevant information but this information was not effectively translated into accurate predictions due to the way the survey elicited forecasts. As this is, to our knowledge, the first study to elicit predictions of client attendance, we encourage future research to explore alternative elicitation methods in larger samples.

4 These barriers to attendance have also been reported in other contexts. For example, a qualitative study with HIV-infected adults in the United States reported stigma, unreliable transport, and competing life activities among the reasons for the non-attendance [34].

